# Routine Blood Tests as Predictors of Periodontal Disease Risk Using a Machine Learning Approach

**DOI:** 10.1101/2024.06.10.24308375

**Authors:** Bushra Ahmad, Khaled Saleh, Saleh Alharbi, Y. Natalie Jeong, Athanasios Zavras, Hend Alqaderi

## Abstract

**Background:** Periodontal disease is a prevalent chronic inflammatory condition associated with systemic health complications. Early identification of high-risk individuals is crucial for targeted preventive and therapeutic interventions. In this study, we aimed to develop and evaluate machine learning models using routine blood tests to predict periodontal disease risk. This will help to develop accessible and cost-effective screening tools for early detection in a non-dental setting.

**Methods:** This study utilized data from the 2013-2014 National Health and Nutrition Examination Survey (NHANES), including full-mouth periodontal examinations, demographic variables, and routine blood tests (complete blood count [CBC], lipid profile, liver and kidney function tests). Periodontitis was defined as a binary outcome based on attachment loss and probing depth at interproximal sites. Individuals meeting any of these criteria were classified as having periodontitis. Seven machine learning models were developed and evaluated using precision, recall, and accuracy metrics.

**Results:** The Random Forest Classifier achieved the highest performance, scoring 0.91 in precision, recall, and accuracy for predicting periodontitis. Key predictive features included smoking status, age, education level, gamma glutamyl transferase, albumin, blood urea nitrogen, glucose, phosphorus, creatinine, basophil count, and total calcium levels.

**Conclusion:** This study underscores the promise of machine learning, especially the Random Forest Classifier, in predicting periodontal disease risk using routine blood tests and demographics. The model accurately identified periodontitis cases, with key features revealing the complex relationship between systemic health and periodontal disease. This suggests the potential for developing machine learning-based screening tools for early periodontal disease detection in non-dental settings.

## 1 INTRODUCTION

Periodontal disease, a chronic inflammatory condition affecting tooth-supporting tissues, is a major public health concern with a global prevalence of 20%-50%.[1] Severe periodontal disease, characterized by extensive tissue destruction and tooth loss, has been associated with systemic conditions such as diabetes, cardiovascular disease, and adverse pregnancy outcomes.[2,3] Early identification of high-risk individuals is crucial for targeted preventive and therapeutic interventions to reduce disease burden and complications.

Routine blood tests, such as CBC and comprehensive metabolic panels are widely accessible and provide valuable information about an individual’s overall health status, including inflammation, nutrition, and organ function.[4] In recent studies, researchers have explored routine blood tests as potential biomarkers for various diseases, including periodontal disease.[5] Associations between blood test parameters (e.g., white blood cell count, C-reactive protein, vitamin D levels) and periodontal disease severity have been reported.[6,7] However, most studies have been limited by small sample sizes and focused on a narrow range of blood test parameters.

Machine learning, a subset of artificial intelligence, is a powerful tool for analyzing complex medical data and developing predictive models for disease risk assessment.[8] Machine learning algorithms can identify patterns and relationships in large datasets, enabling the development of accurate and robust predictive models.[9] In periodontal disease, machine learning approaches have been applied to various data types (e.g., clinical, radiographic, salivary biomarkers) to predict disease progression and treatment outcomes.[10,11] However, the application of machine learning to routine blood test data for predicting periodontal disease risk has not been sufficiently explored.

Given the well-established evidence linking systemic conditions, especially metabolic and inflammatory conditions, to the risk of periodontal disease,[12] it is plausible to hypothesize that certain blood biomarkers may accurately predict the risk of periodontal disease.

In this study, we aimed to develop and evaluate machine learning models using routine blood test data from the NHANES adult population to predict periodontal disease risk. By identifying key blood test parameters associated with periodontal disease and assessing the predictive performance of machine learning models, this study contributes to the development of accessible and cost-effective screening tools for early detection in a non-dental setting.

The NHANES is a nationally representative survey conducted by the Centers for Disease Control and Prevention (CDC) that collects comprehensive health and nutrition data from the US population.[13] The NHANES dataset includes demographic, clinical, and laboratory data, making it a valuable resource for investigating associations between routine blood tests and periodontal disease risk in a large, diverse population.[13]

## 2 MATERIALS AND METHODS

### Study Population

The 2013-2014 NHANES, a cross-sectional survey and publicly accessible survey, provided the data for this study. We complied with the Data Use Restrictions established by the National Center for Health Statistics, and the CDC. The survey employs a complex, multistage, stratified, and clustered sampling design to represent the non-institutionalized civilian US population. NHANES includes a questionnaire, laboratory assays, and clinical examination measures to assess health outcomes and explanatory variables.

The NHANES database indicates that roughly 5,000 individuals, covering a wide age range, participated in at-home interviews and comprehensive health assessments conducted at Mobile Examination Centers. For participants 30 years and older, the survey included full-mouth periodontal examinations performed by pre-calibrated dental professionals within the mobile examination facilities.

### Periodontal Examination

In the NHANES study conducted from 2013 to 2014, participants aged 30 years and older underwent a comprehensive full-mouth periodontal examination (FMPE), excluding those with health conditions requiring antibiotic prophylaxis before periodontal assessment. The primary aim of the FMPE was to obtain gold-standard measurements of clinical attachment loss (AL). To achieve this, two direct measurements were taken at each site: the distance from the cemento-enamel junction to the free gingival margin (CEJ-FGM) and the probing depth (PD). These measurements were collected at six sites (mesiobuccal, midbuccal, distobuccal, mesiolingual, midlingual, and distolingual) on all teeth, except for the third molars. All measurements were rounded down to the nearest whole millimeter. The clinical AL was then determined by calculating the difference between the CEJ-FGM and PD measurements.

### *Periodontitis* (Output)

In this study, periodontitis was defined as a binary outcome, where individuals were categorized as either having periodontitis or not having periodontitis. The presence of periodontitis was determined by the severity of the condition, which was classified into the three categories of mild, moderate, and severe. Mild periodontitis was defined as having ≥ 2 interproximal sites with a ≥ 3 mm attachment loss (AL) and ≥ 2 interproximal sites with ≥ 4 mm PD, not on the same tooth, or 1 site with ≥ 5 mm PD. Moderate periodontitis was characterized by the presence of two or more interproximal sites with ≥ 4 mm clinical AL, not on the same tooth, or two or more interproximal sites with PD ≥ 5 mm, also not on the same tooth. Severe periodontitis was defined as the presence of two or more interproximal sites with ≥ 6 mm AL, not on the same tooth, and one or more interproximal site(s) with ≥ 5 mm PD. If an individual met the criteria for any of these three categories, they were considered to have periodontitis. Conversely, if an individual did not meet any of the criteria for mild, moderate, or severe periodontitis, they were classified as not having periodontitis.

### Routine Blood Tests (Inputs)

We examined lipid profile (low-density lipoprotein, high-density lipoprotein, triglyceride, and total cholesterol), CBC, and liver and kidney function tests (gamma glutamyl transferase (GGT), creatinine, phosphorus, blood urea nitrogen, creatinine phosphokinase (CPK), alkaline phosphatase, aspartate aminotransferase (AST), and alanine aminotransferase (ALT).

### Demographics and other Input Variables

We classified participants’ smoking status as current smokers if they had smoked at least 100 cigarettes in their lifetime and currently smoked daily or occasionally, and as never smokers if they had never smoked 100 or more cigarettes. Diabetes status was a binary variable (diabetic or non-diabetic) based on participants’ self-reported history of diabetes diagnosis by a health care professional. Participants were divided into four age categories: 30 to 34 years, 35 to 49 years, 50 to 64 years, and 65 years and above. Sex was represented as a binary variable, with participants classified as either male or female. The race variable was categorized into five groups: non-Hispanic White, non-Hispanic Black, Hispanic, non-Hispanic Asian, and others. To represent participants’ financial status, we used the ratio of income to the federal poverty level (FPL), dividing this variable into three categories: less than 138% of the FPL, between 138% and 399% of the FPL, and 400% or more of the FPL. Participants’ education levels were categorized into three groups: those who had not completed high school, those who had graduated from high school or obtained a General Education Development high school equivalency test (GED), and those who had pursued education beyond high school. Finally, participants were classified as "Yes" for "Medical condition" if they reported a diagnosis of arthritis, cancer, or coronary heart disease, and as "No" if they did not report any of these diseases.

### Statistical Analysis

We employed Chi-square tests to compare categorical demographic variables stratified by the presence of periodontal disease. We used T-tests and Mann-Whitney tests to compare the levels of blood biomarkers between subjects with and without periodontal disease, depending on the normality of the data distribution. Machine learning models, including Decision Tree, Tuned Decision Tree, Random Forest, Tuned Random Forest, Adaptive Boosting, Gradient Boosting, and XGBoost Classifiers, were developed and evaluated using precision, recall, and accuracy metrics. The most important predictive features were identified for the best-performing model.

### Machine Learning

For the study, we employed a robust methodology to develop and evaluate machine learning models for predicting periodontal disease. We divided the dataset into a training set (70%) for model development and a test set (30%) for assessing predictive performance.

The output variable was a binary variable whether having periodontal disease or not having periodontal disease, and input variables encompassed 14 variables from the routine blood tests, such as lipid profile, CBC, liver and kidney function tests, demographics, and other covariates. We constructed seven models using the Decision Tree, Random Forest Classifier, and XGBoost machine learning algorithms. To address class imbalance in the dataset, we employed a resampling technique by adjusting the ‘class_weight’ hyperparameter in our models.

Specifically, we set ‘class_weight’ to {0: 0.5, 1: 0.5}, ensuring that each class was treated with equal importance, thereby mitigating bias towards the majority class. Additionally, we set the ‘random_statè parameter to ‘1’ to ensure reproducibility of the results.

For the decision tree models, we utilized GridSearch for hyperparameter tuning to compute the optimum values of hyperparameters. The hyperparameters optimized included the maximum depth of the tree (‘max_depth’), specified in the range of 2 to 7, and the minimum number of samples required at a leaf node (‘min_samples_leaf’), with values of 5, 10, 20, and 25. A split point at any depth was only considered if it left at least ‘min_samples_leaf’ training samples in each of the left and right branches. Additionally, we tuned the criterion used to measure the quality of a split, selecting either "gini" for the Gini impurity or "entropy" for information gain. For the Random Forest Classifier, a bagging algorithm was used with decision trees as the base models. Samples were taken from the training data, and each sample’s prediction was made by a decision tree. The final prediction was determined by combining the results from all the decision trees through voting. The hyperparameters tuned for the Random Forest Classifier included the number of trees in the forest (‘n_estimators’), the minimum number of samples required to split an internal node (‘min_samples_split’), with values of 1, 2, 5, and 10, the minimum number of samples required at a leaf node (‘min_samples_leaf’), with values of 1, 4, and 10, and the number of features to consider when looking for the best split (‘max_features’), with values of 0.5 and ‘auto’. We employed XGBoost (Extreme Gradient Boosting), a tree-based ensemble machine learning technique, for model building. Hyperparameter tuning for boosting included algorithms such as Adaboost, Gradient Boosting, and XGBoost. This involved optimizing parameters to enhance model performance and ensure robust predictive capabilities. The model performance was evaluated using recall, precision, and accuracy. These metrics were calculated from the confusion matrix. Recall measures the model’s ability to correctly identify positive instances, precision measures the accuracy of the positive predictions made, and accuracy provides the overall correctness of the model. To identify the most influential features in predicting periodontal disease, we conducted a feature importance analysis on the best-performing model. We used Stata 17 for descriptive analysis and employed Python for machine learning analysis.

## 3 RESULTS

Table 1 presents the descriptive characteristics of the study population, stratified by periodontitis status. Significant differences were observed between the two groups in terms of various demographic and health-related factors. Participants with periodontitis were more likely to be current smokers, male, older, and of Hispanic or Black race/ethnicity than those without periodontitis. They also tended to have lower educational attainment and lower income levels. Additionally, diabetes was more prevalent among participants with periodontitis. However, no significant difference was found between the two groups in terms of the presence of any medical condition.

**TABLE 1:**
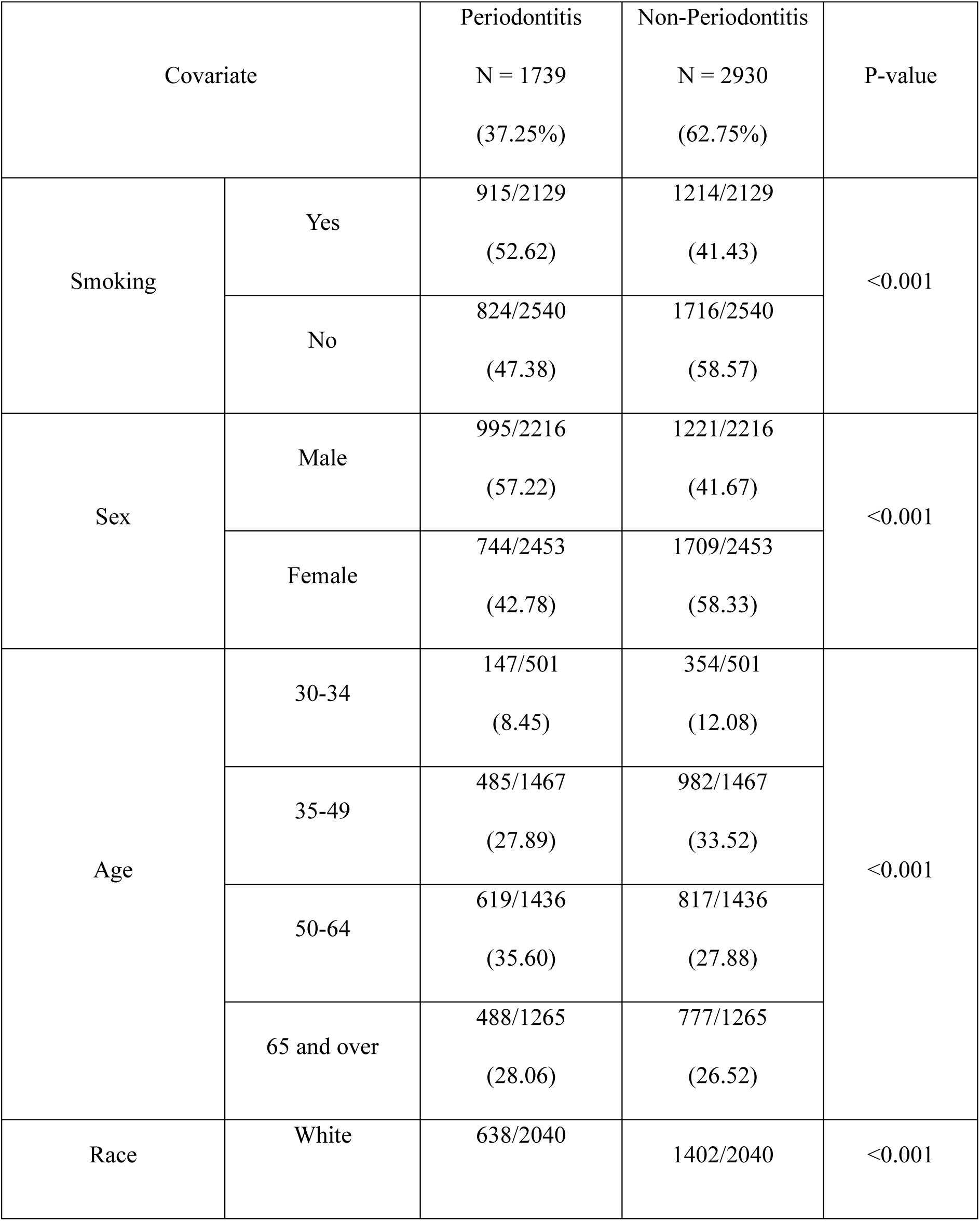
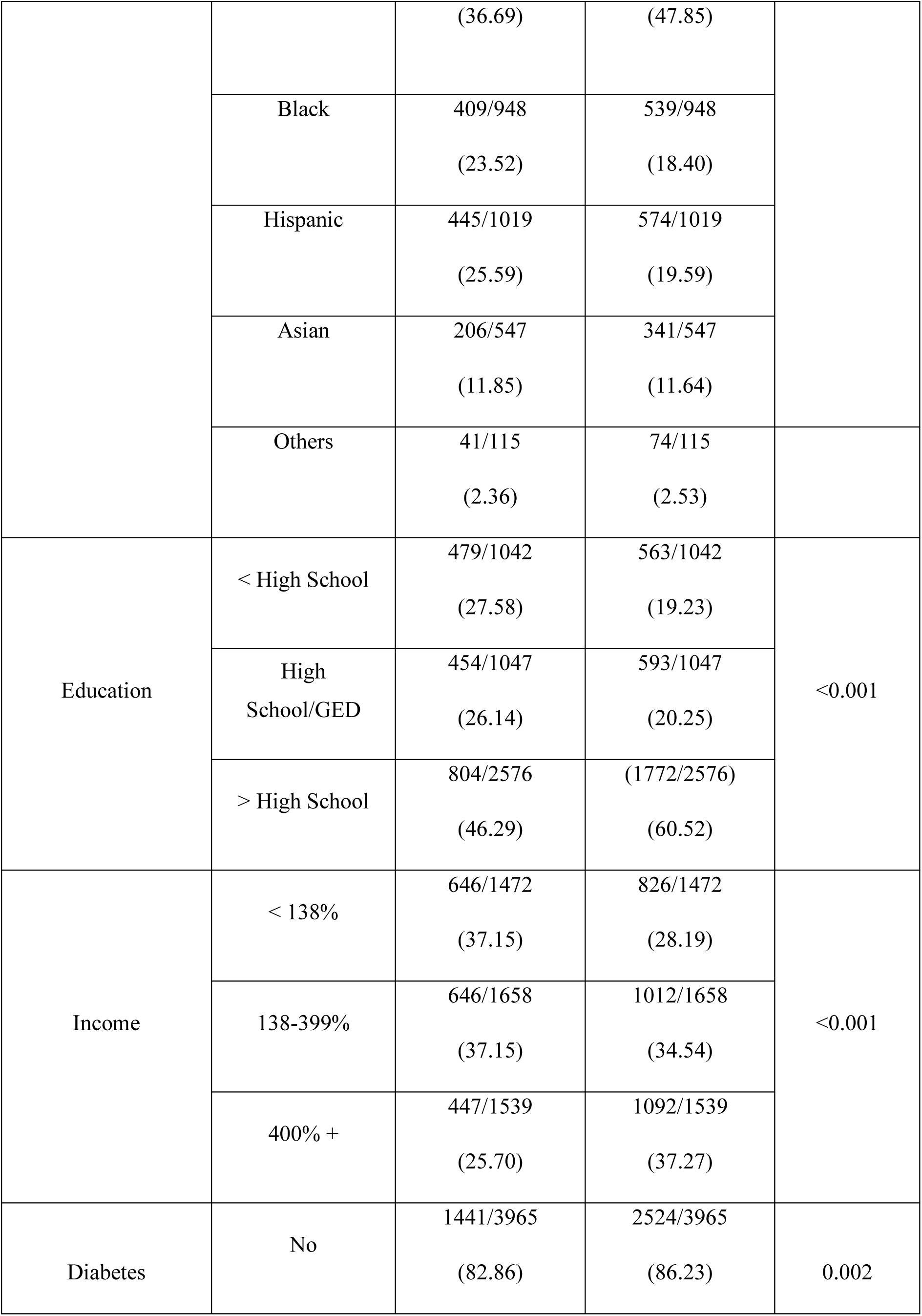
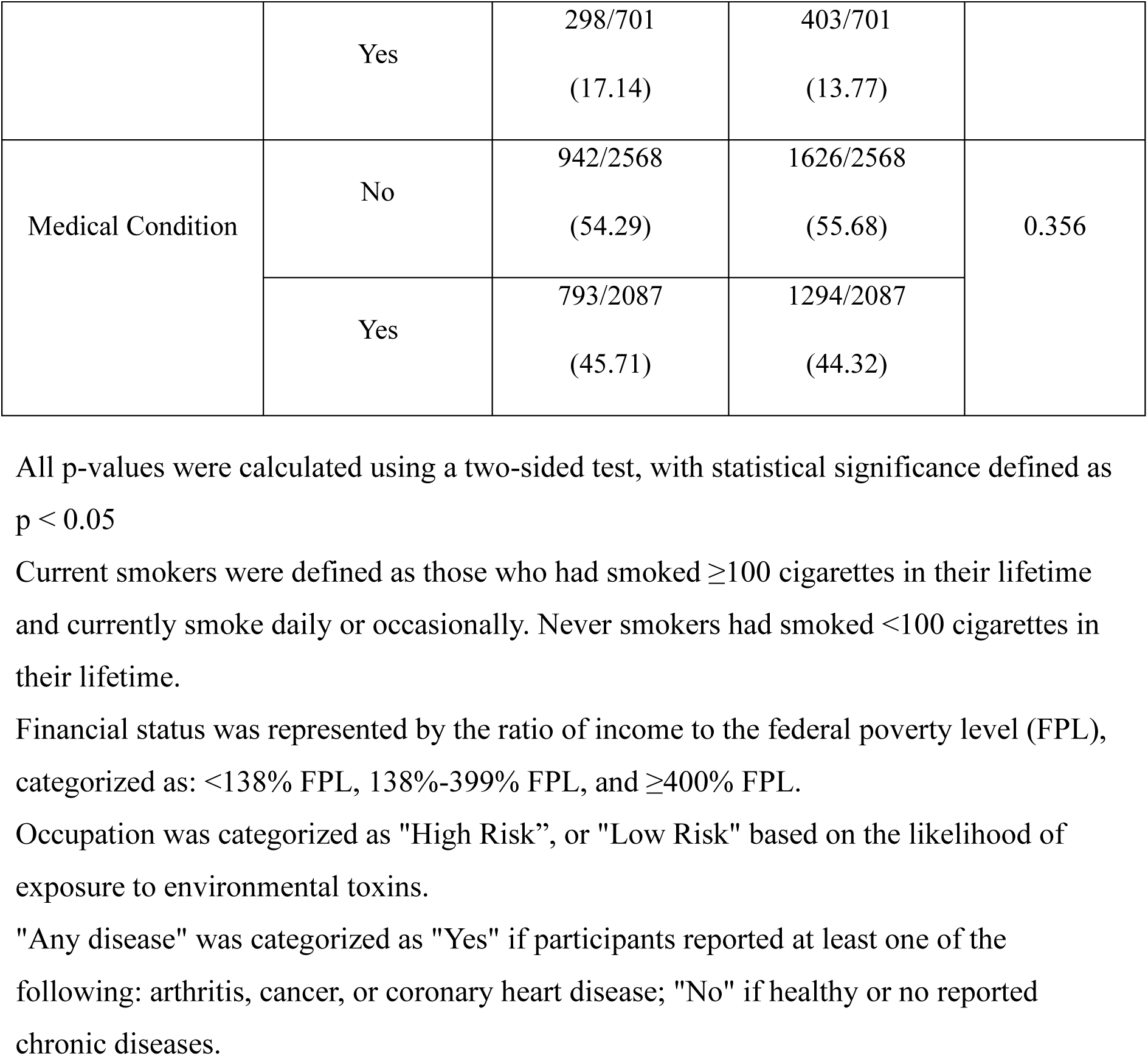
Descriptive summary of the population characteristics with and without periodontitis.

Table 2 summarizes the levels of various blood biomarkers in patients with and without periodontitis. Patients with periodontitis exhibited significantly higher levels of glucose, creatinine, gamma glutamyl transferase, and basophil number compared to those without periodontitis. In contrast, albumin and phosphorus levels were significantly lower in the periodontitis group. No significant differences were observed in blood urea nitrogen and total calcium levels between the two groups.

**TABLE 2:** Descriptive summary of blood biomarkers in patients with and without periodontitis.

| Blood Biomarkers | No Periodontitis<br>Median (IQR) | Periodontitis<br>Median (IQR) | P-value |
| --- | --- | --- | --- |
| Glucose refrigerated serum (mg/dL) | 95 (19) | 98 (23) | <0.001† |
| Creatinine (mg/dL) | 0.86 (0.30) | 0.88 (0.31) | 0.005† |
| Blood urea nitrogen (mg/dL) | 13 (6) | 13 (6) | 0.2607† |
| Gamma glutamyl transferase (U/L) | 19 (15) | 21 (17) | <0.001† |
| Basophil number (1000 cells/uL) | 0 (0.1) | 0.1 (0.1) | 0.0087† |
| Albumin (g/dL)<br>Mean (SD) | 4.23 (0.33) | 4.20 (0.32) | 0.0067* |
| Phosphorus (mg/dL)<br>Mean (SD) | 3.82 (0.59) | 3.76 (0.59) | 0.0021* |
| Total calcium (mg/dL)<br>Mean (SD) | 9.44 (0.37) | 9.44 (0.38) | 0.8184* |

\*T-test
†Mann-Whitney

Table 3 compares the performance of various machine learning models in predicting periodontitis. The Random Forest Classifier demonstrated the highest performance across all three metrics, scoring 0.91 across precision, recall, and accuracy. Conversely, the Tuned Decision Tree Classifier scored the last with 0.80 across the board.

**TABLE 3:**
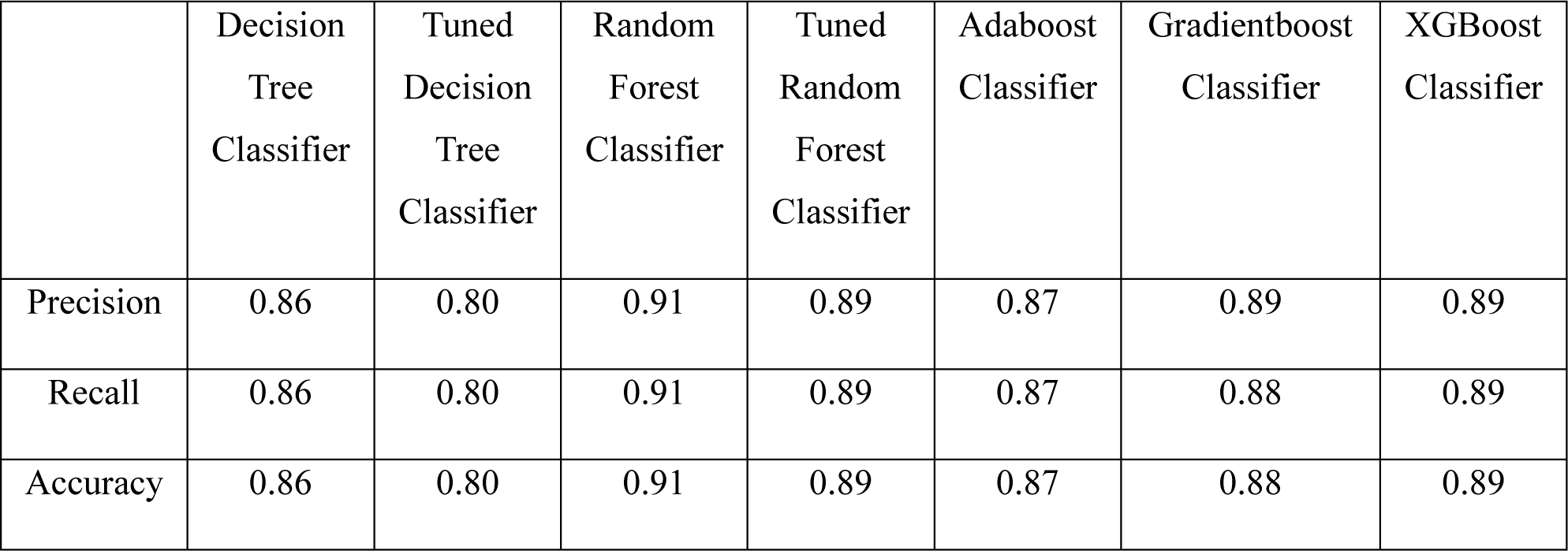
Comparison of the performance of various machine learning models in predicting periodontitis.

|  | Decision<br>Tree<br>Classifier | Tuned<br>Decision<br>Tree<br>Classifier | Random<br>Forest<br>Classifier | Tuned<br>Random<br>Forest<br>Classifier | Adaboost<br>Classifier | Gradientboost<br>Classifier | XGBoost<br>Classifier |
| --- | --- | --- | --- | --- | --- | --- | --- |
| Precision | 0.86 | 0.80 | 0.91 | 0.89 | 0.87 | 0.89 | 0.89 |
| Recall | 0.86 | 0.80 | 0.91 | 0.89 | 0.87 | 0.88 | 0.89 |
| Accuracy | 0.86 | 0.80 | 0.91 | 0.89 | 0.87 | 0.88 | 0.89 |

Figure 1 shows the confusion matrix for the best-performing model which details the true positives (91%), false negatives (9%), false positives (10%), and true negatives (90%) produced by the model.

**FIGURE 1:**
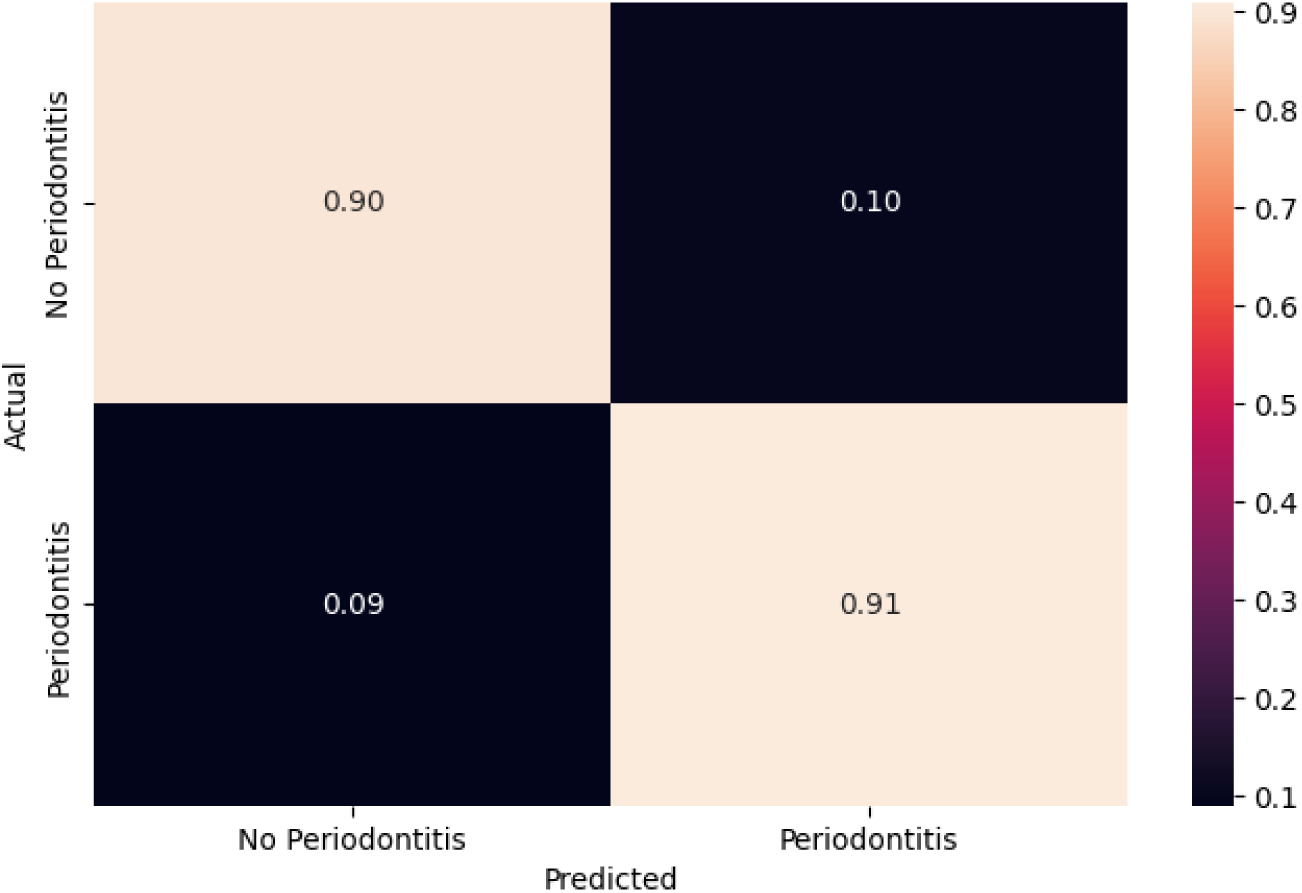
Confusion matrix of the Random Forest. Classifier model for predicting periodontitis.

Figure 2 demonstrates the most important features that predict periodontitis. The features included demographic factors such as education level, age, sex, and income. Smoking, CBC tests, and liver and kidney function tests were also included.

**FIGURE 2:**
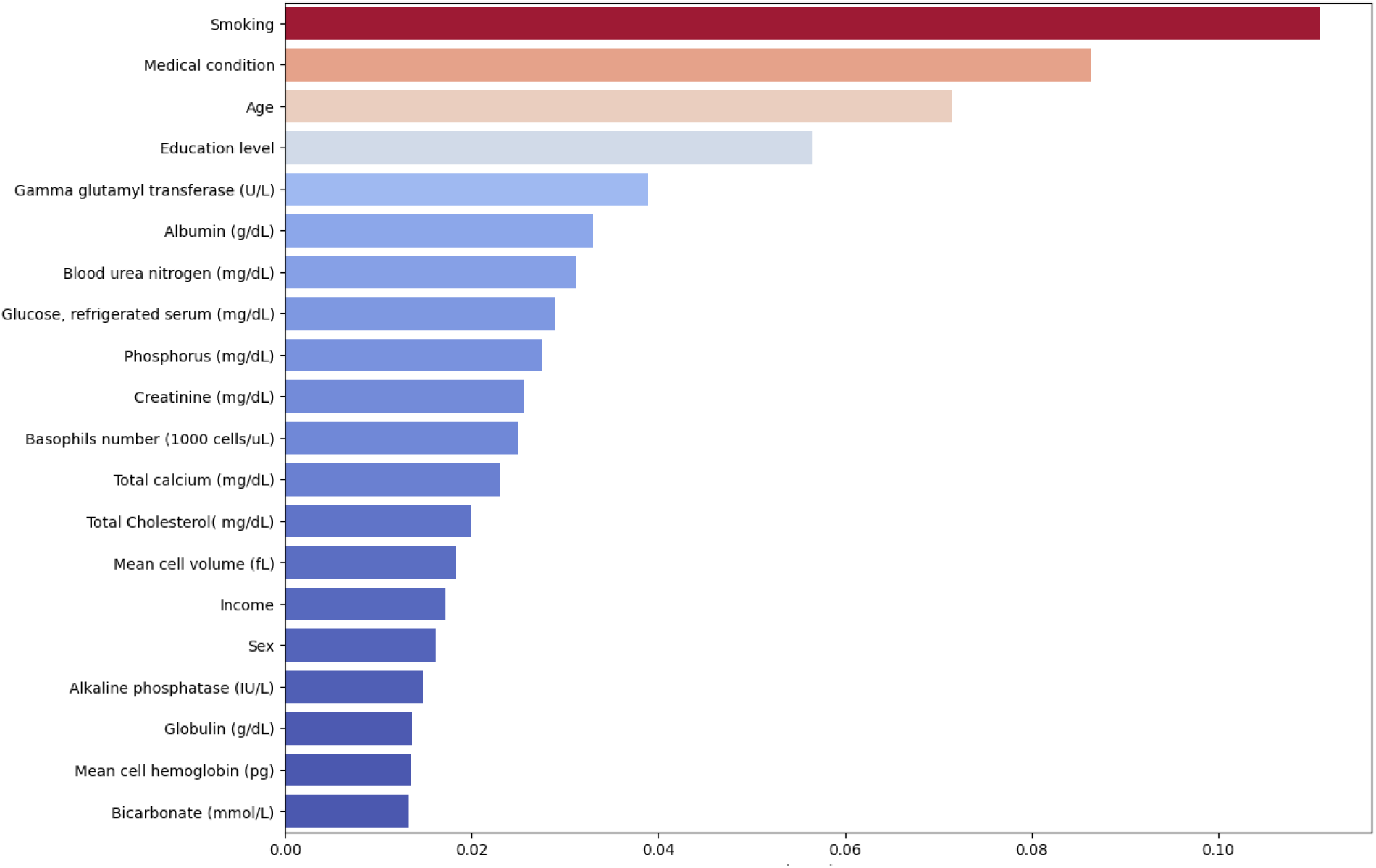
Feature importance ranking for periodontitis prediction model.

## 4 DISCUSSION

We evaluated seven machine learning models using routine blood test data from a representative sample of the US adult population to predict the risk of periodontal disease. All models demonstrated high accuracy, precision, and recall, ranging from 80% to 91%. The Random Forest Classifier achieved the highest performance, with precision, recall, and accuracy of 0.91. These results indicate that routine blood test measurements combined with demographic factors can accurately predict periodontal disease risk.

Smoking status emerged as the strongest predictive feature in our study, with current smokers being at higher risk for periodontitis compared to non-smokers. This finding is consistent with the well-established evidence that smoking is a major risk factor for periodontal disease.[14] Smoking has been shown to impair the immune response, alter the oral microbiome, and compromise the healing capacity of periodontal tissues, leading to increased susceptibility to periodontal destruction.[15]

Age was another important predictive feature in our study, with older individuals being at higher risk for periodontal disease. This finding is supported by numerous epidemiological studies that have demonstrated an increased prevalence and severity of periodontal disease with advancing age.[16] The cumulative exposure to risk factors over time, along with age-related changes in the immune system and tissue homeostasis, may contribute to the higher risk of periodontal disease in older adults.[17]

Education level was also identified as one of the top predictive features, which is in line with research consistently showing a strong association between lower education levels and higher prevalence and severity of periodontal disease.[18] Individuals with lower education levels may have limited access to dental care, and lower oral health literacy, and may engage in health-compromising behaviors, all of which contribute to an increased risk of periodontal disease.[19]

GGT, a liver enzyme associated with oxidative stress and systemic inflammation, has been reported to have elevated levels in patients with periodontal disease, indicating a link between liver function and periodontal health.[20,21] Sanikop et al. found that GGT levels were significantly higher in patients with chronic periodontitis compared to healthy controls and positively correlated with the severity of periodontal destruction.[22] Our findings further support the association between GGT and periodontal disease, highlighting its potential as a biomarker for periodontal risk assessment.

Furthermore, serum albumin was another predictive feature in our model. Serum albumin is the main protein synthesized by the liver and is a practical marker of general health status, demonstrating the severity of underlying diseases. Inflammation and malnutrition both reduce albumin concentration by decreasing its rate of synthesis.[23] Ogawa et al. suggest an inverse relationship between periodontal disease and serum albumin concentration, possibly influenced by nutritional aspects and inflammatory markers such as C-reactive protein.[23] The decreased serum albumin levels associated with periodontitis may be attributed to the impairment of the host response, including phagocytic function, cell-mediated immunity, complement system, secretory antibody, and cytokine production and function, potentially contributing to the progression of periodontal destruction.[23]

Blood urea nitrogen (BUN) is a marker of kidney function. Various factors can influence BUN levels, such as kidney function, protein intake, and catabolic states.[24] Our model revealed that BUN played a role in predicting periodontitis, in addition to other variables. Some studies have reported associations between elevated BUN and periodontal disease severity. Kshirsagar et al. found that individuals with periodontal disease had a higher prevalence of renal insufficiency, as indicated by elevated BUN levels.[24] Akar et al. reported that BUN levels were significantly higher in patients with chronic periodontitis than in healthy controls.[25] The evidence suggests a link between impaired kidney function, reflected by elevated BUN, and periodontal disease progression, possibly due to the bidirectional relationship between these two conditions.

Blood glucose was one of the most influential features for predicting periodontitis in our study, aligning with the well-known bidirectional relationship between hyperglycemia or diabetes and periodontal disease. Individuals with diabetes have a higher risk of periodontitis, and periodontal inflammation can induce insulin resistance and worsen glycemic control.[26] Hyperglycemia contributes to an exaggerated inflammatory response, impaired wound healing, altered oral microbial composition, amplifying periodontal destruction.[27] Our results, combined with substantial evidence, emphasize the importance of screening for diabetes and monitoring glycemic status in individuals at high risk for periodontitis to enable timely intervention and co-management.

Our study identified serum phosphorus levels as a significant predictive feature, with lower levels associated with increased periodontitis risk, aligning with previous research suggesting an inverse relationship between serum phosphorus and periodontal disease.[28] Although the underlying mechanisms remain unclear, they may involve phosphorus’s role in bone metabolism and inflammation.[28] Further research is needed to elucidate the precise relationship.

Creatinine, a marker of kidney function, was identified as a predictor of periodontal disease in our study.[29] Previous research by Arora et al. and Kapellas et al. supports this association, demonstrating higher creatinine levels in patients with chronic periodontitis and linking elevated creatinine with an increased risk of periodontal disease.[30,31] Our findings reinforce this link and suggest that creatinine may be a useful biomarker for periodontal risk assessment.

Our study identified basophil count as an important predictive feature of periodontal disease. Recent studies support this finding, suggesting a potential role of basophils in the pathogenesis of periodontal disease. Basophils release histamine and other inflammatory mediators, contributing to the inflammatory process in periodontal tissues.[32] Arun et al. found a significant increase in basophil count in patients with chronic periodontitis compared to healthy controls.[33] Agarwal et al. reported that basophil count was positively associated with the severity of periodontal disease.[34] These findings, along with our results, highlight the potential of basophil count as a biomarker for periodontal disease risk assessment.

Our study identified total calcium levels as an important variable in predicting the risk of periodontitis. Machado et al. noted that the relationship between serum calcium levels and periodontitis remains unclear.[35] However, other studies have reported conflicting results, suggesting a potential link between altered calcium homeostasis and periodontal disease.[36,37] Various factors, including dietary intake, hormonal regulation, and certain medical conditions, can affect serum calcium levels.[35] Although the exact mechanisms linking calcium to periodontitis remain unclear, alterations in calcium metabolism may influence periodontal health through its effects on bone metabolism, immune function, or other physiological processes.[35] Further research exploring the relationship between calcium homeostasis, periodontal disease, and other systemic factors could clarify calcium’s role in the pathogenesis of periodontitis.

Artificial intelligence and machine learning techniques have the potential to revolutionize periodontal disease risk assessment and prediction.[8, 38] These technologies utilize advanced statistical modeling and algorithmic learning to uncover complex relationships within large datasets, enabling the development of highly accurate prediction tools.[39,40] Recent studies have demonstrated promising applications in periodontal disease management, including predicting treatment outcomes, detecting disease from radiographs, and identifying high-risk individuals.[10,41,42] Our findings further highlight the importance of integrating medical and dental care by demonstrating that routine blood tests can predict the risk of developing periodontitis. This machine learning model provides a valuable tool for early detection and prevention of periodontitis in primary care and non-dental healthcare settings.

Based on our findings, we have several recommendations. We propose that medical professionals, particularly those in primary care, incorporate periodontal risk assessments into their evaluations using accessible blood parameters. To effectively implement these recommendations, we suggest developing a standardized screening protocol that uses our predictive model to identify individuals at high risk for periodontitis. Once identified, these patients can be referred to dental professionals for further evaluation and treatment, fostering a collaborative approach between medical and dental care providers.

By bridging the gap between these two domains of healthcare, we can enhance the early detection and management of periodontitis, ultimately leading to improved oral and overall health outcomes for patients. This integrated approach can also raise awareness among medical professionals about the importance of oral health and its connection to systemic well-being, encouraging a more comprehensive and holistic view of patient care.

While our study offers valuable insights, it is limited by its cross-sectional design, preventing the assessment of temporal relationships, and its reliance on a single NHANES cycle, potentially limiting generalizability. Future research should validate these models in independent cohorts and explore incorporating additional clinical and biological markers alongside routine blood tests to enhance prediction accuracy. Prospective studies are also needed to examine the temporal relationships between predictive factors and disease progression.

## 5 CONCLUSION

In conclusion, our study demonstrates the potential of machine learning models to accurately predict periodontal disease risk using routine blood tests and demographic data. Identifying key biomarkers like GGT, albumin, BUN, glucose, phosphorus, creatinine, basophil count, and total calcium, our model offers a valuable tool for early detection and risk assessment in non-dental settings. Integrating this into routine medical exams could facilitate timely referrals and proactive management, improving oral and overall health. Further research is needed to validate these findings in diverse populations and explore additional biomarkers for enhanced predictive accuracy of machine learning models for periodontal disease risk assessment.

## Data Availability

The data that supports the findings of this study are available from the corresponding author upon reasonable request.

## AUTHOR CONTRIBUTIONS

Bushra Ahmad designed the study, acquired data, assisted with data analysis, and wrote the manuscript. Khaled Saleh contributed to the writing of the manuscript, interpreting the data, and revising the manuscript. Saleh Alharbi revised the manuscript. Hend Alqaderi supervised the study, assisted with machine learning analysis, and revised the manuscript. Y. Natalie Jeong provided critical revisions of the manuscript. All authors approved the final version of the manuscript to be published and agreed to be accountable for all aspects of the work.

## Notes

### Competing Interest Statement

The authors have declared no competing interest.

### Funding Statement

This research did not receive any specific grant from funding agencies in the public, commercial, or not-for-profit sectors.

